# Why does left ventricular distension usually not occur in veno-arterial extracorporeal membrane oxygenation?

**DOI:** 10.1101/2022.08.02.22278301

**Authors:** Manoj Myneni, Faisal Cheema, Keshava Rajagopal

## Abstract

**Objectives:** Using a lumped parameter-based theoretical circulatory model, we sought to examine the potential effects of veno-arterial (V-A) extracorporeal membrane oxygenation (ECMO) support on coronary blood flow and resultant left ventricular (LV) contractility.

**Background:** Previous theoretical studies have suggested that V-A ECMO support results in markedly increased left ventricular intracavity pressures and volumes, i.e., not only inadequate unloading, but exacerbated loading. However, this phenomenon of LV distension occurs only uncommonly in clinical scenarios. This discrepancy between previous theoretical work and clinical experience requires explanation.

**Methods:** We defined a piecewise linear relation between the end-systolic pressure and volume of the ventricles to simulate ascending and descending limbs of the Frank-Starling relationship. A linear relationship between coronary blood flow rate and left ventricular contractility was added, defining the so-called “Gregg effect”.

**Results:** LV systolic dysfunction resulted in reduced coronary blood flow; V-A ECMO support augmented coronary blood flow, proportionally to the circuit flow rate. On V-A ECMO support, a weak or absent Gregg effect resulted in increased LV end-diastolic pressures and volumes, and increased end-systolic volume with decreased LV ejection fraction (LVEF), consistent with LV distension. In contrast, a more robust Gregg effect resulted in unaffected and/or even reduced LV end-diastolic pressure and volume, end-systolic volume, and unaffected or even increased LVEF.

**Conclusions:** In this lumped parameter model-based theoretical study, V-A ECMO support was found to augment coronary arterial blood flow. A resultant proportional augmentation of LV contractility may be an important contributory mechanism underlying why LV distension is uncommon in the setting of V-A ECMO support.

## Introduction

Cardiogenic shock is an important cause of mortality and morbidity attributed to cardiovascular disease. Left ventricular (LV) failure is the most common physiological subtype of cardiogenic shock (1), and in turn, acute coronary syndromes constitute the most common cause of acute left ventricular failure (2). Short-term mechanical circulatory support (MCS) is a critically important component of the treatment of cardiogenic shock, along with treatment of the underlying cause(s) if feasible (3).

Veno-arterial (V-A) extracorporeal membrane oxygenation (ECMO) is commonly used to provide short-term MCS in the setting of cardiogenic shock in general (4), and LV failure in particular. The advantages of V-A ECMO most notably are: 1) provision of partial or complete cardiac and pulmonary function, and 2) ease/rapidity of the institution, without requirements of image guidance for cannulation.

However, MCS may not be **cardiac-beneficial** support. Cardiac-beneficial support is thought to relate to ventricular *unloading*, i.e., a reduction in the ventricular loading conditions (most commonly construed in lumped parameter models as “preload” and “afterload”). Unloading is thought to be salubrious to cardiomyocytes because myocardial oxygen consumption is known to be proportional to and causally linked to “pressure-volume area” (PVA), the sum of the stroke work (SW) and what is commonly termed “potential energy” (PE; we will refer to this as unavailable potential energy (UPE) since it is PE that *cannot* be converted into work/kinetic energy under the specific conditions examined) (5).

### Hypothesis and Purpose

Unlike other MCS modalities that involve left/systemic ventricular (LV), and to a lesser extent even left atrial (LA) circuit inflow/drainage, V-A ECMO does not directly drain the LV. Previous theoretical studies suggested that LV unloading is not only not achieved, but that LV intracavity pressures (by virtue of V-A ECMO flow into the systemic arterial circulation, for a fixed systemic arterial impedance, thereby augmenting LV afterload) and volumes are actually augmented (6-8); diastolic pressure/volume augmentation, particularly with reduced contractility, constitutes LV distension. Thus V-A ECMO is commonly not thought to unload the LV or be beneficial for it.

Yet, clinical experience is substantially different from these theoretical studies. Only a minority of patients develop LV distension following V-A ECMO (< 30% overall, and < 10% with clinically important as opposed to hemodynamic distension (9)). We have previously discussed the conceptual/theoretical bases underlying LV distension in this setting (10), and suggested some as yet unidentified mechanism(s) that are operational in preventing the occurrence of LV distension. This suggests that one or both of the following is true: 1) important assumptions about the “initial conditions” to which the models are applied are wrong, and/or 2) the models themselves, and their constitutive/governing equations, are either incomplete or wrong.

One possible mechanism contributing to the lack of LV distension in most patients with LV failure supported via V-A ECMO is an increase in LV contractility and trans-aortic valve LV ejection due to V-A ECMO-induced augmentation of coronary blood flow. The generalized phenomenon of coronary blood flow augmentation-caused increase in LV contractility, most noteworthy when coronary blood flow increases from a previously low level at which LV dysfunction had occurred, was first observed and reported by Gregg in the 1950s (11). We hypothesized that the Gregg effect might explain why LV distension is relatively uncommonly observed in V-A ECMO support, and sought to test this in a theoretical study.

## Methods

### Cardiovascular system model

A lumped parameter electrical analog model for the cardiovascular system proposed by Ursino (12) was modified and extended by adding a coronary circulation originating from the arterial side of the systemic circulation and emptying into the right atrium (RA). Briefly, the model consists of systemic and pulmonary circulations, each modeled using resistances, compliances, and an inertance, and a time-varying elastance model for the right and the left ventricles. The RA and LA were modeled as capacitances. A schematic of the circuit (Figure S1) and the equations governing systemic and pulmonary circulations are given in the supplementary material. The pressure in the LV (*P*_*lv*_) was calculated by solving the following equation

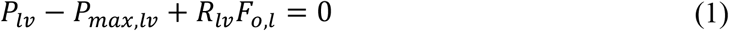

where *P*_*max,lv*_ is the pressure in the LV if the cardiac cycle were to be nonejecting (isovolumic) and defined through the equations below using a time-varying elastance model

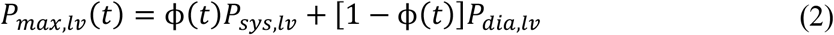

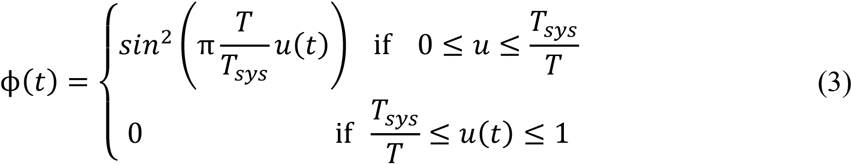

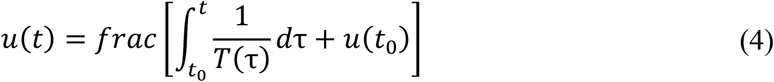

where *T* is the heart period and *T*_*sys*_ is the duration of systole. *frac* function resets the value of *u*(*t*) to 0 once it reaches the value 1. We adopted the piecewise linear function proposed by Wang et al. (13) to approximate the unimodal end-systolic pressure-volume relationship (ESPVR)

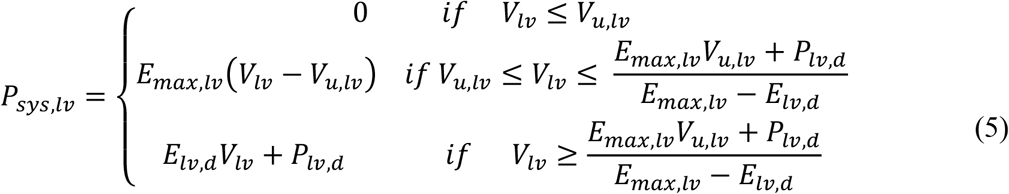

An exponential relation was used to represent the end-diastolic pressure-volume relation

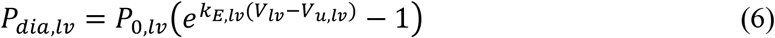

The rate of the ejection of blood from the LV into systemic circulation was defined as

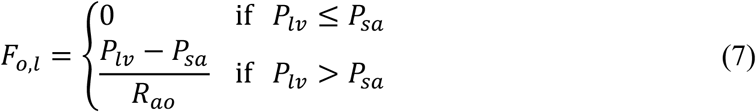

where *R*_*ao*_ is the resistance of the aortic valve. *R*_*lv*_ is the viscous resistance that signifies the reduction of maximum pressure generated by the LV due to the ejection of blood compared to an isovolumic cardiac cycle. It was assumed to be linearly related to the maximum pressure during an isovolumic cardiac cycle.

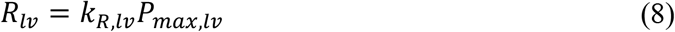

Similar equations were used to model the right ventricle. Thus the *first* fundamental difference between our model and others (notably that of Burkhoff and coworkers (6)) is a unimodal ESPVR that captures ventricular distension-related ventricular dysfunction.

### Coronary circulation model with Gregg effect

The coronary circulation was represented using the model proposed by Bovendeerd et al (14). The model defines an intramyocardial pressure (*p*_*im*_), which is a function of the LV pressure and the LV volume, and represents the coronary circulation using constant resistances and compliances. Governing equations for the model are given in the supplementary material.

To account for the influence of the coronary blood flow rate on the contraction of the LV (Gregg effect), we define two values of the mean coronary blood flow rate over one cardiac cycle (*Q*_*c*_): *Q*_*c,max*_ and *Q*_*c,min*_ between which *E*_*max,lv*_ and *Q*_*c*_ are linearly related. Beyond *Q*_*c,max*_, an increase in *Q*_*c*_ has a negligible effect on *E*_*max,lv*_.

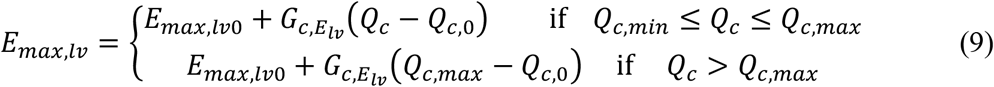

where *E*_*max,lv*0_ and *Q*_*c*,0_ are the values of *E*_*max,lv*_ and *Q*_*c*_ in the absence of any mechanical circulatory support, *Q*_*c,min*_ is the mean flow rate below which *E*_*max,lv*_ drops rapidly with a reduction in *Q*_*c*_. To calculate *Q*_*c*_ at any time *t*, the coronary blood flow rate is averaged over the time interval [*t* − 5*T*, t]. Thus, the *second* key difference between our newly modified model and others is the inclusion of the Gregg effect.

### V-A ECMO circuit

Circulatory support provided by V-A ECMO was simulated by taking the inflow from the RA to the ECMO circuit and giving the outflow from the ECMO circuit to systemic arteries. We limit ourselves to the overall effect of V-A ECMO in pumping blood from the systemic venous to the systemic arterial side by fixing the blood flow rate between 0-5 L/min. This approximation was sufficient to study the response of the cardiovascular system to V-A ECMO support.

### Systolic LV failure

Systolic LV failure was simulated by reducing the value of *E*_*max,lv*_. *P*_*lv,d*_ was defined to vary linearly with *E*_*max,lv*_, indirectly reflecting the reduction in the maximum pressure a failing LV can generate.

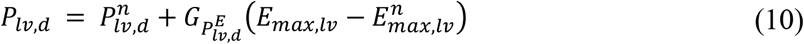

where 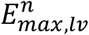 and 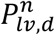 represent the values of the slope of ascending limb of the ESPVR and *P*_*lv,d*_, respectively for a normal left ventricle.

## Results

### Hemodynamic response to V-A ECMO support in the absence of coronary blood flow-ventricular contractility interaction

Figure 1 shows the variation of hemodynamic variables with V-A ECMO flow rate in the absence of coronary blood flow-ventricular contractility interaction. With increasing severity of LV failure, a reduction in systemic mean arterial pressure (MAP) was observed, the extent of this decrease being proportional to the severity of LV failure, accompanied by similar increases in MPAP, LVEDP, and LVEDV. Circulatory support provided by V-A ECMO flow improved MAP and reduced MPAP at all levels of LV failure. A reduction in systemic arterial pulse pressure was also observed, the extent of this decrease being proportional to the ECMO flow rate, at all levels of LV failure. A significant increase in LVESV (and ESP – Figure 2) was observed in a fashion proportional to the ECMO flow rate. LVEDV (and EDP) also increased similarly, although by a smaller amount compared to LVESV (and ESP). Consequently, LVEF decreased as the ECMO flow rate increased for normal as well as failing LVs. The increase in LVEDV as a function of ECMO flow rate was higher when LV failure was more severe. For moderately 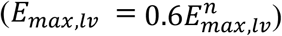 and severely 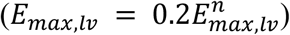 failing LV, LVSW decreased as ECMO flow rate increased. For mild LV failure 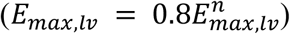, however, LVSW remained nearly constant. LVPVA increased as a function of ECMO flow rate for all cases of LV failure, consistent with an increase in the oxygen consumption of the left ventricle. However, coronary perfusion also improved with ECMO support, in a manner proportional to the ECMO flow rate.

**Figure 1:**
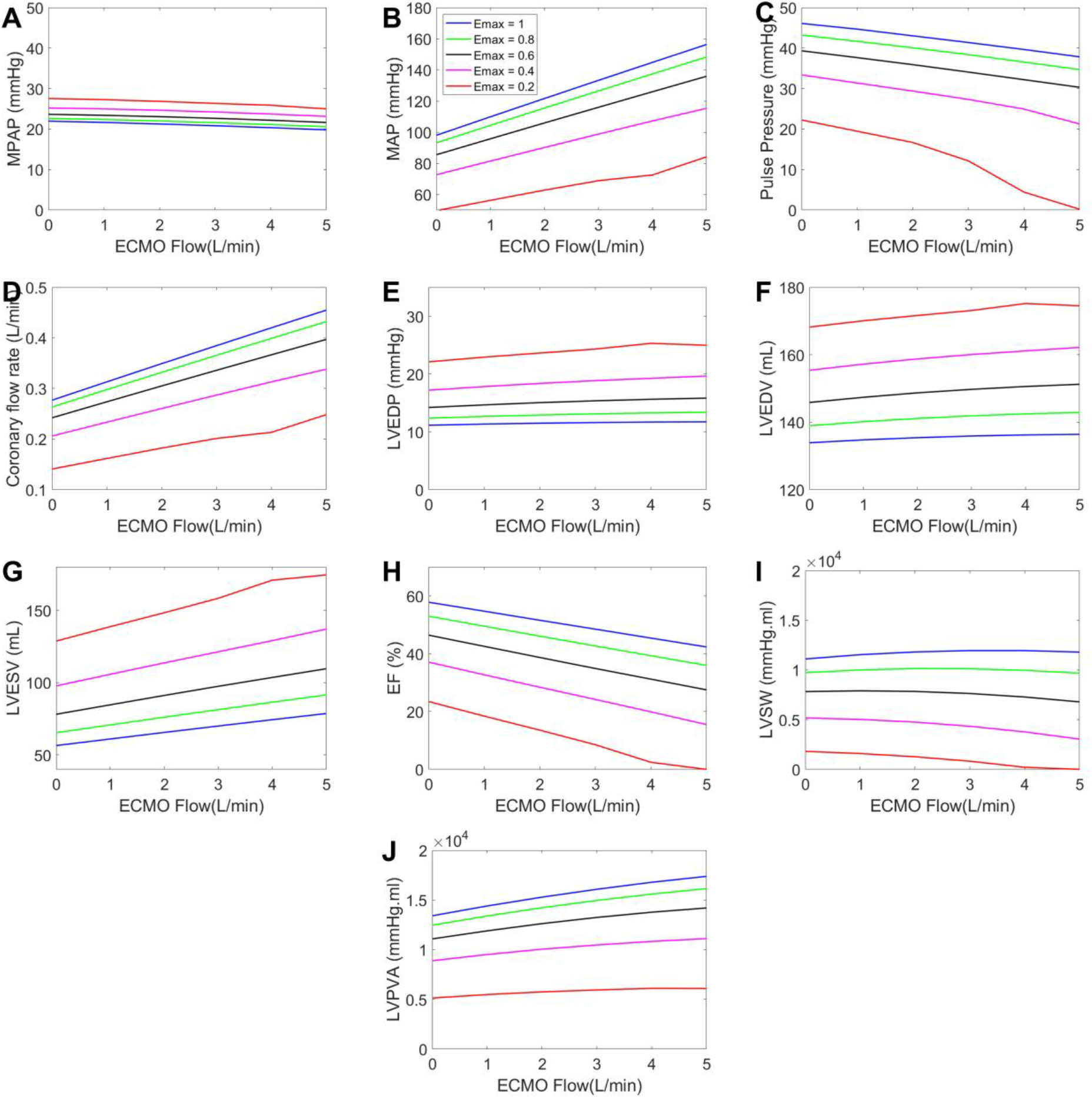
Hemodynamic response without the Gregg effect. Variation of hemodynamic variables with ECMO (extracorporeal membrane oxygenation) flow rate in the absence of coronary-ventricular interaction (i.e., when the Gregg effect is not considered). Legend represents the ratio 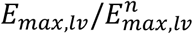. MPAP – mean pulmonary arterial pressure; MAP – mean systemic arterial pressure; LV – left ventricle; EDP - end-diastolic pressure; EDV - end-diastolic volume; ESV - end-systolic volume; EF – LV ejection fraction; SW – stroke work; PVA – pressure-volume area.

**Figure 2:**
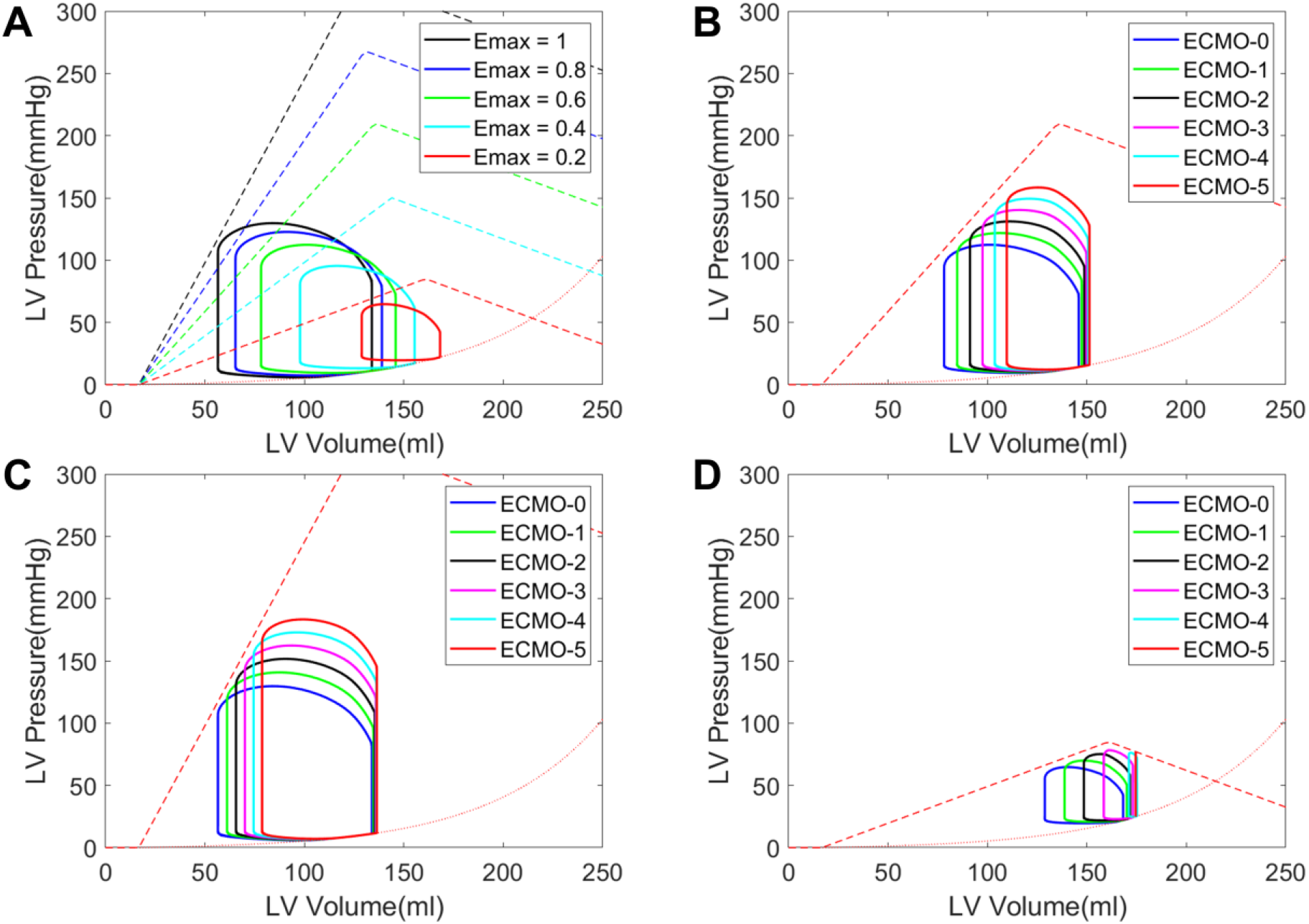
LV PV diagrams without the Gregg effect. LV PV diagrams in the absence of the Gregg effect. (a) Increasing severity of heart failure with no circulatory support, legend represents the ratio 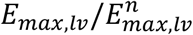 (b) A normal left ventricle with increasing ECMO flow rate, (c) Moderate left ventricular failure with 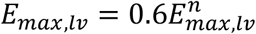 at different ECMO flow rates, (d) Severe left ventricular failure with 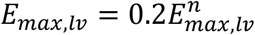 at different ECMO flow rates. Legend represents V-A ECMO (veno-arterial extracorporeal membrane oxygenation) flow rate in L/min. LV- left ventricle; PV- pressure-volume.

Figure 2 shows LV PV diagrams without circulatory support and with varying levels of LV failure (Figure 2A), and at different levels of ECMO flow rate for normal (Figure 2B), moderately failing (Figure 2C), and severely failing LVs (Figure 2D). A rightward shift of the PV loop, with increased LVEDV and LVESV, was observed with increased severity of LV failure (Figure 2A). V-A ECMO support further increased LVESV (and ESP, consistent with an increased afterload, in the absence of any changes in LV contractility) and LVEDV in the cases of moderate and severe LV failure. Furthermore, in severe left ventricular failure (figure 2D), increasing ECMO flow rate results in progressive LV dilation along with a reduction in contractility (descending limb of the ESPVR) with increasing EDV and EDP, these last two of which define LV distension. At 5 L/min ECMO flow rate, the left ventricle dilates considerably, and the aortic valve is closed (i.e., pulse pressure is 0). Thus, with severe pre-existing LV systolic dysfunction, as ECMO flow rate increases, distension occurs, with exacerbated LV dysfunction manifesting in progression to the descending limb of the Frank-Starling relationship.

### Hemodynamic response to V-A ECMO support: Influence of coronary blood flow-ventricular contractility interaction

As shown, coronary perfusion improved as ECMO flow rate increased. We considered, as a function of coronary blood flow, an improvement in the contractility index (*E*_*max,lv*_) of the left ventricle (Gregg effect). These results are shown in Figure 3 for normal, moderately failing, and severely failing LVs. The values of 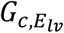and *Q*_*c,max*_ for these simulations were taken to be 3 × 10^−3^ (mmHg/ml)/(ml/min) and 300 ml/min, respectively. When this effect was considered, MAP and coronary blood flow rate increased with increasing ECMO flow rate, and MPAP decreased with increasing ECMO flow rate, as in the absence of the Gregg effect. In moderate LV failure, the arterial pulse pressure decreased with an increase in the ECMO flow rate. In contrast, in severe LV failure, the arterial pulse pressure was constant (minimally increased in fact) with ECMO flow rates of up to 4 L/min. LVEDP and LVEDV were essentially constant for moderate LV failure. Interestingly, in severe LV failure, LVEDP and LVEDV decreased modestly with ECMO flow rates of up to 4 L/min; this is in contrast to the increase observed with the absence of the Gregg effect (Figure 3E vs. Figure 1E and Figure 3F vs. Figure 1F, respectively). That is, for a given ECMO flow rate, LVEDV (and EDP) is lower in the presence of the Gregg effect. For a given ECMO flow rate, a substantial reduction in LVESV was observed compared to the case when the Gregg effect was not considered (Figure 3G vs. Figure 1G). The reduction in ejection fraction was lesser compared to the case without the Gregg effect (Figure 3H vs. Figure 1H). For moderate LV failure, LVSW increased up to 2 L/min ECMO flow rate. LVSW increased as a function of ECMO flow rate until 4 L/min for severe LV failure. LVPVA also increased, some of which is due to the increase in LVSW, with increasing ECMO flow rate. In Figure 4, LV PV diagrams are shown for moderate and severe LV failure cases with the Gregg effect (Figure 4A and 4B), and for the severe LV failure case at ECMO flow rates of 2 L/min (Figure 4C) and 4 L/min (Figure 4D) with and without the Gregg effect. In the presence of the Gregg effect, the PV loop of the LV shifted towards lower LVEDV (and EDP) and LVESV (but higher ESP due to increased LV contractility). Notably, in severe LV failure, in the absence of the Gregg effect 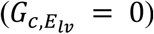, the PV loop of the LV is in the descending limb of the ESPVR at ECMO flow rate of 4 L/min; this is an exhibition of true LV distension (i.e., dilatation-exacerbated/induced LV systolic dysfunction) for the case of severe LV failure. In the presence of the Gregg effect, however, the PV loop of LV remained in the ascending portion of the ESPVR.

**Figure 3:**
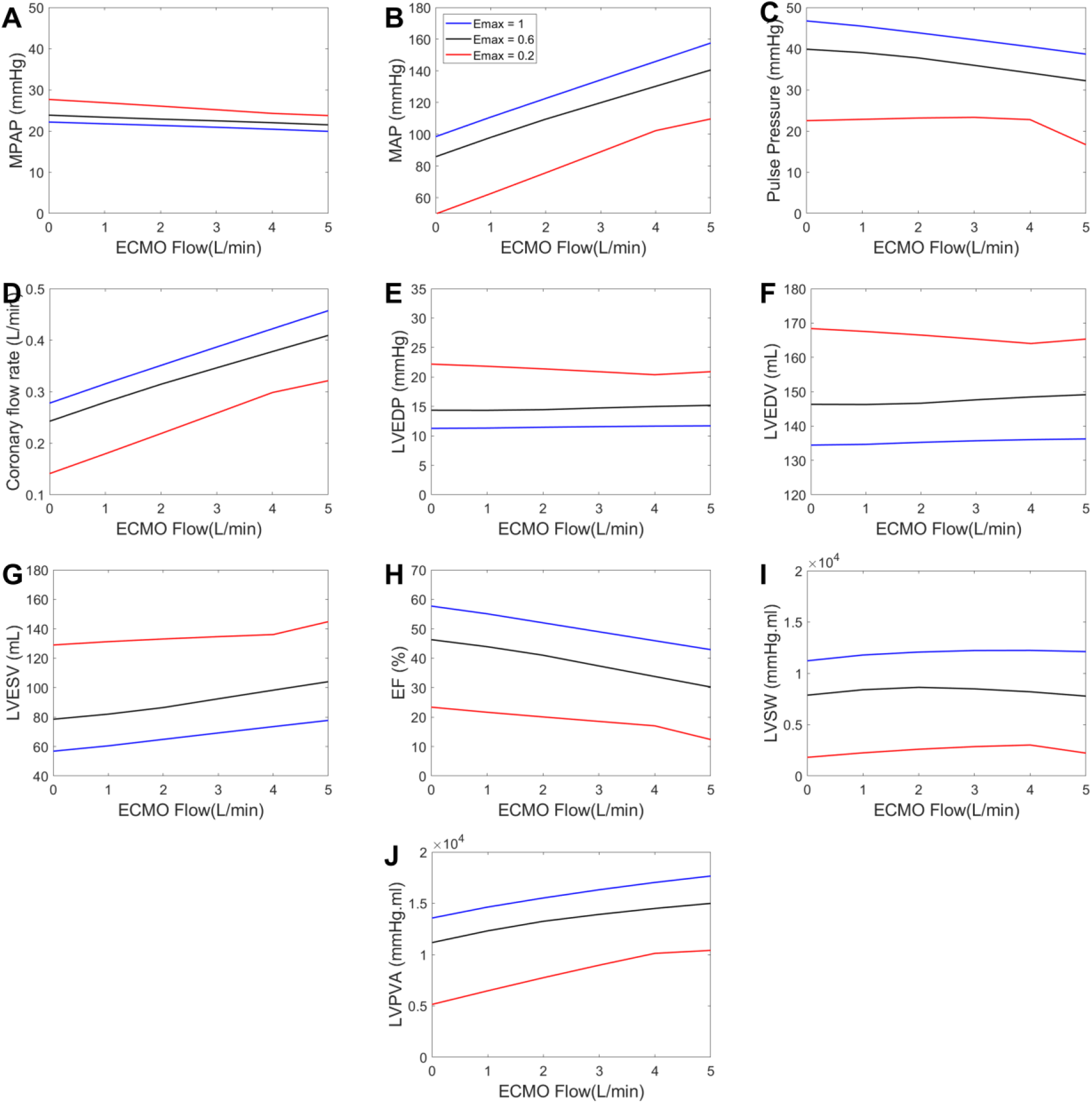
Hemodynamic response with the Gregg effect. Variation of hemodynamic variables with ECMO flow rate when the Gregg effect is considered. Legend represents the ratio 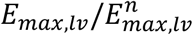. The values of 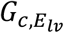 and *Q*_*c,max*_ are 3 × 10^−3^ (*mmHg/ ml*)*/*(*ml/min*) and 300 ml/min, respectively. MPAP – mean pulmonary arterial pressure; MAP – mean systemic arterial pressure; LV – left ventricle; EDP - end-diastolic pressure; EDV - end-diastolic volume; ESV - end-systolic volume; EF – LV ejection fraction; SW – stroke work; PVA – pressure-volume area.

**Figure 4:**
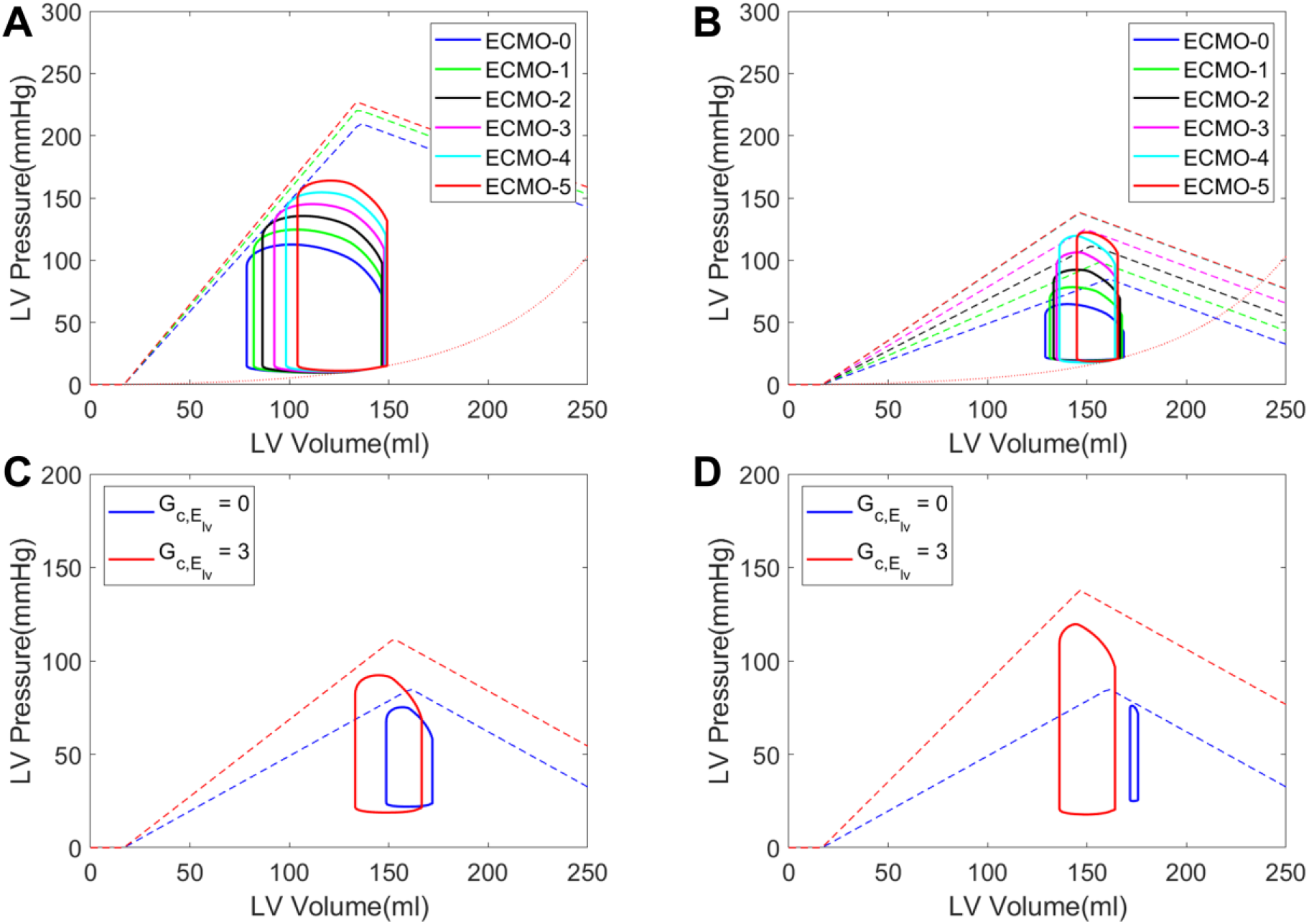
LV PV diagrams with the Gregg effect. LV PV diagrams for (A) Moderate left ventricular failure, 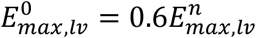 with increasing ECMO (extracorporeal membrane oxygenation) flow rate (legend in L/min) and the Gregg effect taken into account, (B) Severe left ventricular failure 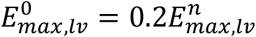 with increasing ECMO flow rate (legend in L/min) and the Gregg effect taken into account, (C) With and without the Gregg effect at ECMO flow rate of 2L/min in the case of severe LV failure, (D) With and without Gregg effect at ECMO flow rate of 4L/min in the case of severe LV failure. The values of 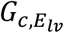 and *Q*_*c,max*_ for the cases with the Gregg effect are 3 × 10^−3^ *(mmHg/ml)/(ml/min)* and 300 ml/min, respectively. LV- left ventricle; PV- pressure-volume.

### Parametric Study

#### Influence of Q_c,max_ on the hemodynamic response

The effect of changes in *Q*_*c,max*_ (maximal coronary flow rate above which, *E*_*max,lv*_ no longer changes with changes in *Q*_*c*_) on the hemodynamic response under V-A ECMO was found to be relatively modest and the details are presented in the supplementary material.

#### Influence of the magnitude of the Gregg effect on the hemodynamic response

We analyzed the consequences of changing 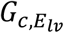, which indicates the magnitude of the Gregg effect coupling coronary blood flow to LV contractility, between 0 − 6 × 10^−3^ (mmHg/ml)/(ml/min), on the hemodynamic response for the case of severe LV failure. For these simulations, the value of *Q*_*c,max*_ was fixed at 300 ml/min. Figure 5 presents the variation of hemodynamic variables with ECMO flow rate for different 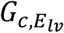 in the setting of severe LV failure. MAP increases and MPAP decreased as a function of ECMO flow rate for all values of 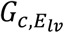. For smaller values of 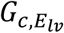 (i.e., absent or a “weak” Gregg effect), LVEDV (and EDP) increased proportionally to ECMO flow rate, indicating LV dilation. In contrast, at larger values (≥ 3 × 10^−3^ (mmHg/ml)/(ml/min)), which characterize a “stronger” Gregg effect, LV unloading occurred with a reduction in LVEDV up until the ECMO flow rate above which ventricular contractility no longer increases. The optimal ECMO flow rate at which the maximum LV unloading occurs depends on the strength of the Gregg effect. At a fixed ECMO flow rate, a higher value of 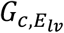 resulted in a larger reduction in LVEDV, thereby providing larger LV unloading. Even LVEF improved at larger values of 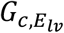, with a large reduction in LVESV. LVSW increased as ECMO flow rate increased until it reaches a maximum, for 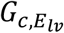 values of 3 × 10^−3^ (mmHg/ml)/(ml/min) and beyond, whereas for smaller 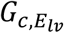 values, LVSW decreased as ECMO flow rate increased. The unavailable potential energy (UPE) increased with the ECMO flow rate. LVPVA increased as a function of ECMO flow rate, and the change in LVPVA from its value in the absence of ECMO support was larger when the Gregg effect was stronger. We observe that the fraction of the LVPVA that is accounted for by LVSW (Figure 5J), which reflects LV mechanical efficiency increased with an increase in the value of 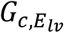, showing the beneficial effect that V-A ECMO flow has on LV function as the magnitude of the Gregg effect increased. Accompanying this increase in LVSW is a reduction in the *fraction* of LVUPE (Figure 5K) with an increase in the value of 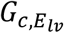, again consistent with the Gregg effect and improved LV contractility. The ratio Δ*LVPVA*/Δ*Q*_*c*_ (which may be regarded as the ratio of change in energetic demand to change in supply; Figure 5L) also decreased with increasing ECMO flow rate for larger values of 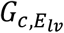 (≥ 3 × 10^−3^ (mmHg/ml)/(ml/min)), indicating that the increased LV energetic demand on V-A ECMO (Δ*LVPVA*) was accompanied and more than compensated by the V-A ECMO-induced augmentation in coronary blood flow (Δ*Q*_*c*_) when the Gregg effect is robust.

**Figure 5:**
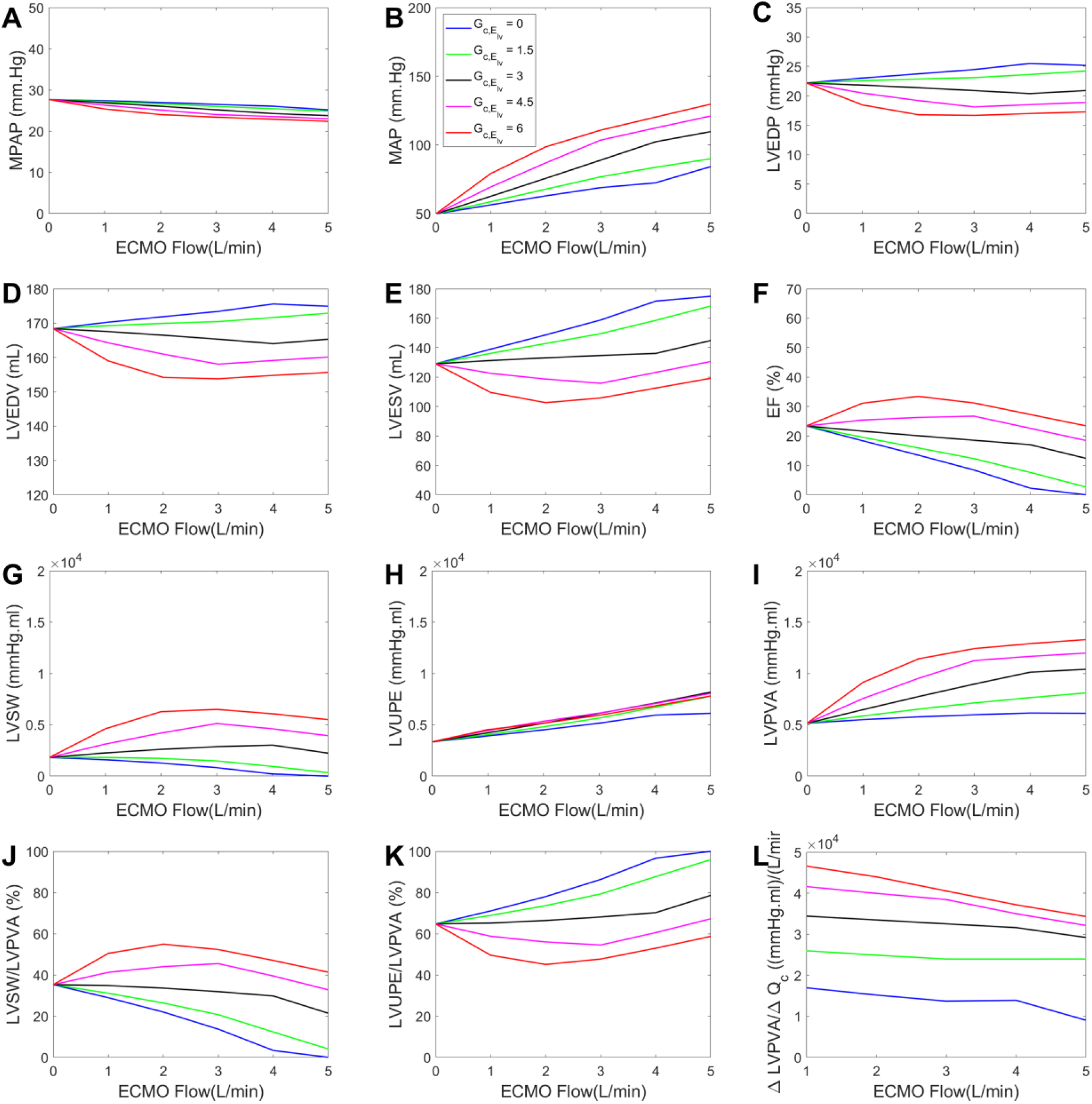
Influence of 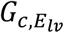 (magnitude of the Gregg effect) on hemodynamic response. Variation of hemodynamic variables with ECMO (extracorporeal membrane oxygenation) flow rate for different 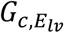 in the case of severe left ventricular failure 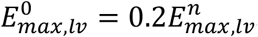. The value of *Q*_*c,max*_ is fixed at 300 ml/min. Legend represents the value of 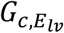 in × 10^−3^ *(mmHg/ml)/(ml/min)*. Δ*A* is the difference between the value of the variable *A* at a given ECMO flow rate and its value in the absence (0 L/min) of ECMO support. MPAP – mean pulmonary arterial pressure; MAP – mean systemic arterial pressure; LV – left ventricle; EDP - end-diastolic pressure; EDV - end-diastolic volume; ESV - end-systolic volume; EF – LV ejection fraction; SW – stroke work; UPE – unavailable potential energy; PVA – pressure-volume area.

In summary, in severe LV systolic dysfunction, as coronary perfusion improves under V-A ECMO support, a “stronger” Gregg effect results in LV unloading, with a nearly constant or reduced LVEDV and LVEDP, and/or LVESV, increase in LVSW as well as its ratio to LVPVA, signifying an improvement in LV systolic function under ECMO support. Arterial pulse pressure also increases under ECMO support in the presence of a “stronger” Gregg effect. On the other hand, an absent or a “weaker” Gregg effect results in LV dilation, reduction in LVSW and its ratio to LVPVA, and an increase in the ratio of LVUPE to LVPVA, indicating the detrimental effect V-A ECMO may have on LV function when the Gregg effect is weak or absent (Central Illustration).

## Discussion

In this theoretical study, we sought to ask and answer (at least *in silico*) the question: why does left ventricular distension usually *not* occur in the setting of V-A ECMO support? V-A ECMO is overwhelmingly most commonly used to treat cardiogenic shock; in turn, cardiogenic shock overwhelmingly most commonly is characterized by severe LV systolic dysfunction (3, 4). Our circulatory model differs from those used in the bulk of previous studies in two key ways, the second of which is unique to our knowledge. First, we utilize a unimodal ESPVR, which more faithfully captures the actual phenomenon of LV distension, wherein high EDV and EDP actually *cause* exacerbated LV systolic dysfunction/contractility reduction. Second, in hypothesizing that a proportional relationship between coronary blood flow and LV contractility (Gregg effect) may play an important role in explaining the infrequency of LV distension, we incorporate the Gregg relationship into our model.

We found that: (1) LV systolic failure results in reduced coronary blood flow, which V-A ECMO support in turn improves, consistent with previously published experimental data (15, 16), (2) in the absence of the Gregg effect, V-A ECMO support is associated (within the model, causally) with LV distension, the extent of which is proportional to the severity of LV dysfunction as well as the ECMO flow rate, and (3) in the presence of the Gregg effect, V-A ECMO is not associated with LV distension, and with a robust Gregg effect, the LV becomes unloaded.

These findings may explain at least in large part why V-A ECMO usually does not result in LV distension. Previous theoretical studies have predicted the consistent occurrence of LV distension following V-A ECMO support (6-8), which is inconsistent with clinical experience (9). Moreover, although such studies employ lumped parameter models that are incapable of distinguishing between spatial variations in variables (for example, central versus peripheral cannulation), it is generally appreciated that central V-A ECMO cannulation is less likely to result in LV distension unless either a systemic-to-pulmonary artery shunt or aortic regurgitation is present, and that when LV distension does occur, it is in the setting of peripheral cannulation (9). This is particularly relevant to cardiac transplantation undertaken in the context of donation after cardiac determination of death (DCDD) and thoracoabdominal normothermic regional perfusion (TA-NRP) (17). In the DCDD with TA-NRP, V-A ECMO with central cannulation (albeit typically with a reservoir, which typically is present in cardiopulmonary bypass but not in ECMO circuits (10) is used to resuscitate the donor heart following donor withdrawal of care and progression to normothermic cardiac arrest. The basis of resumption of cardiac electrical and mechanical activity is extrinsically (via V-A ECMO) derived coronary blood flow. Although TA-NRP in DCDD is different from typical V-A ECMO (the latter usually with peripheral cannulation and without a reservoir), it highlights the physiological and clinical importance of ECMO circuit-driven coronary blood flow and the recovery of ventricular systolic function.

With respect to previous experimental work, one of the early studies on hemodynamic effects of V-A ECMO support on post-ischemic hearts (warm ischemia) of sheep by Bavaria et al.(18) observed a slight reduction in LVEDV and LVESV with increasing ECMO flow. They further reported an improvement in LV contractility due to ECMO support, and suggested improved coronary perfusion as a possible mechanism for improved contractility. Our model demonstrated a reduction in LVESV and LVEDV with ECMO flow rate compared to baseline for severe LV failure when 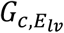 is 4.5 × 10^−3^ and 6 × 10^−3^(mmHg/ml)/(ml/min). Similar to our results for the cases with 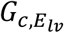 4.5 × 10^−3^ and 6 × 10^−3^ (mmHg/ml)/(ml/min) in severe LV failure, Brehm et al. (16) reported that V-A ECMO support resulted in an increase in the blood flow through the left anterior descending coronary artery and a reduction in the LA pressure (or equivalently LVEDV or LVEDP) in pigs suffering from cardiogenic shock induced by esmolol infusion. More recently, Ostadal et al. (19) studied the effects of V-A ECMO support in a swine model of heart failure created by coronary artery occlusion using a balloon catheter and introducing deoxygenated blood downstream of the balloon. They observed no statistically significant changes in LV EDV, increased LVESV, and a decreased ejection fraction as a function of the ECMO flow rate. We observed similar changes in LVESV, LVEDV, and LVEF in the case of severe heart failure when 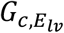 is 3 × 10^−3^ (mmHg/ml)/(ml/min) (Figure 3). In their study, LVSW increased up to 4L/min ECMO flow rate and decreased slightly thereafter. Our simulations depicted the same trend in LVSW for the case of severe heart failure when 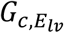 of 3 × 10^−3^ (mmHg/ml)/(ml/min) (Figure 3G). Kawashima et al. (20) studied heart failure in dogs induced by sequential ligation of LAD coronary artery branches. For moderate and severe failure cases, they observed PVA to increase with ECMO support. Our simulations show the same, irrespective of the influence of the Gregg effect. However, in their experiments, PVA decreased on V-A ECMO for mild cases of LV dysfunction, which we did not observe in our simulations.

### Study limitations

The phasic nature of the blood flow in the coronary circulation is modeled using an intramyocardial pressure that is a function of LV volume and LV pressure. The resistances and capacitances of the coronary circulation are assumed to be constant as a function of time during the cardiac cycle, during which LV pressure and volume of course vary. In reality, this is incorrect, and time-varying impedance of the coronary circulation ideally ought to be incorporated (21, 22). Next, we have modeled the ESPVR using a continuous non-differentiable piecewise linear function; in contrast, the actual ESPVR is a continuous and differentiable curve. We have not considered the effect of baroregulation on the hemodynamic response during V-A ECMO support. We have not simulated the effects of V-A ECMO in case of chronic cardiomyopathies and these findings may not directly translate to these settings. Finally, a limitation inherent in any lumped parameter model is the absence of any *spatial* variation in hemodynamic variables. As discussed previously, the model presented here cannot differentiate between peripheral and central cannulation.

Despite these limitations, based upon our review of the literature, this appears to be the first study of its kind, in evaluating the potential impact of the effects of V-A ECMO support upon coronary blood flow and LV contractility/systolic function, and the consequent presence or absence of LV distension. Future theoretical studies will focus on providing additional modeling complexity in order to address the aforementioned limitations.

## Conclusions

In this lumped parameter model-based theoretical study, V-A ECMO support was found to augment coronary arterial blood flow in the setting of LV failure. V-A ECMO support was observed to be associated with LV distension in the absence of the Gregg effect. In contrast, V-A ECMO support was found to unload the left ventricle in the presence of a robust Gregg effect (coronary blood flow augmentation-caused increase in LV contractility) and was not associated with LV distension.

## Implications for Clinical Practice

### Competency in medical knowledge

It is well known that myocardial ischemia is causally linked both to acute and chronic left ventricular systolic dysfunction in the context of coronary artery disease. However, the Gregg effect, which characterizes a broad proportional relationship between coronary blood flow and left ventricular contractility, is not well appreciated in the clinical context. The Gregg effect may be an important mechanism underlying the absence of left ventricular distension in most cases of V-A ECMO. In addition, it may be important in understanding cardiac recovery on V-A ECMO and other modalities of short-term mechanical circulatory support.

### Translational Outlook

Both the assessment and optimization of coronary blood flow in the setting of mechanically assisted circulation to treat shock are not well-studied. This theoretical study suggests that further experimental and clinical work focused on coronary blood flow and its regulation may be valuable in improving cardiac function and survival in cardiogenic shock treated via mechanical circulatory support.

## Data Availability

All data produced in the present study are available upon reasonable request to the authors

## Figure titles and legends

**Central Illustration:**
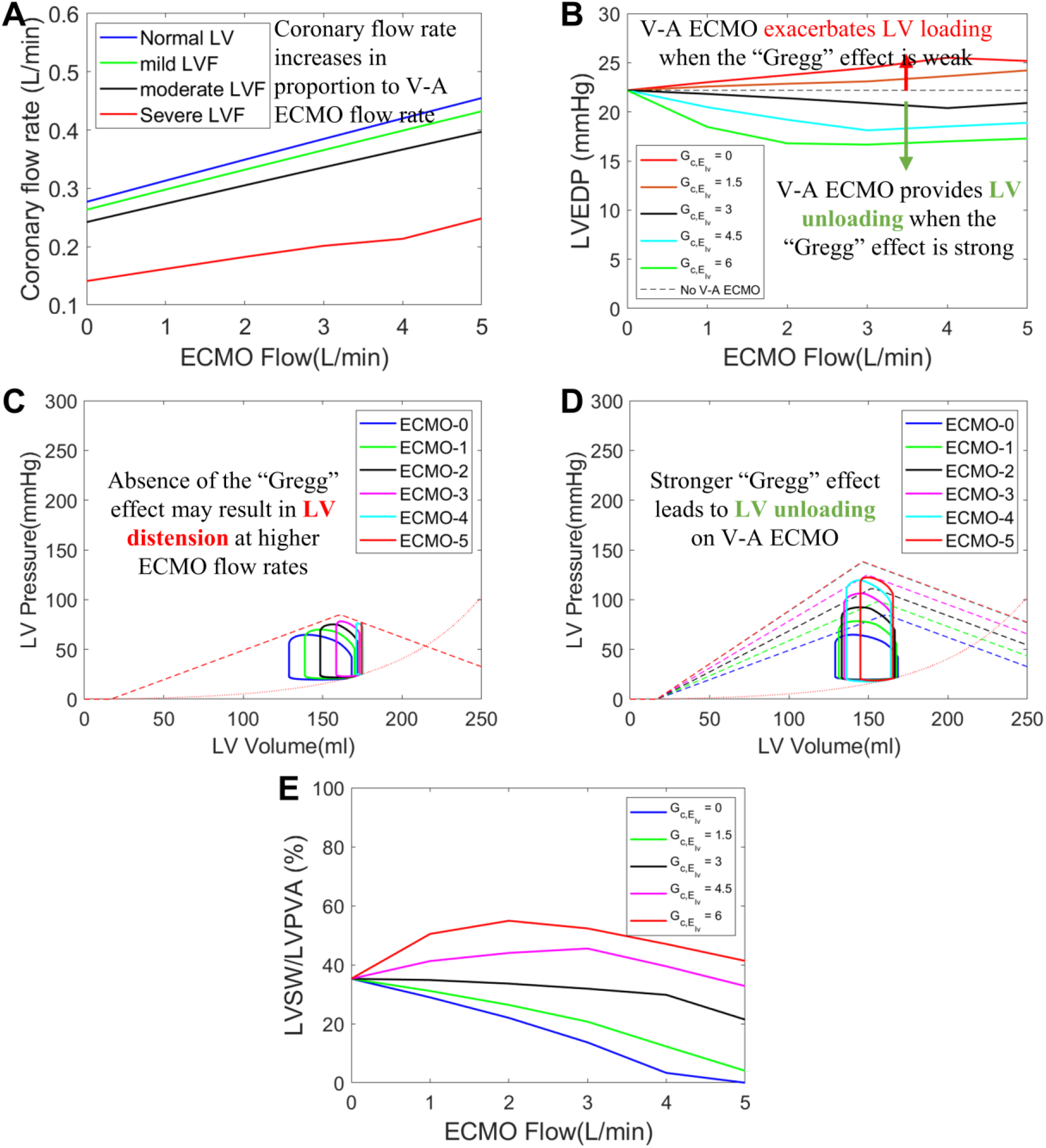
Influence of the Gregg effect on LV loading and function during V-A ECMO support. (A) Coronary flow rate increases proportionally to the V-A ECMO (veno-arterial extracorporeal membrane oxygenation) circuit flow rate (legend shows the severity of LV failure). (B) V-A ECMO unloads LV in the presence of a stronger Gregg effect; LV loading is exacerbated by V-A ECMO when the Gregg effect is weak. (C,D) LV pressure-volume diagrams in the absence and in the presence (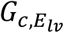 of 3 × 10^−3^ (mmHg/ml)/ (ml/min)) of the Gregg effect, respectively at different levels of V-A ECMO flow rate (legend in L/min). In the absence of the Gregg effect, V-A ECMO support is associated (within the model, causally) with LV distension, the extent of which is proportional to the ECMO flow rate, and in the presence of the Gregg effect, the LV becomes unloaded and V-A ECMO is not associated with LV distension. (E) Mechanical efficiency of LV increases under ECMO support in the presence of a stronger Gregg effect. 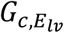 values are shown in × 10^−3^ (mmHg/ml)/ (ml/min). LV-left ventricle.

## Abbreviations and Acronyms

EDV: end-diastolic volume
EDP: end-diastolic pressure
EF: ejection fraction
ESPVR: end-systolic pressure-volume relationship
ESV: end-systolic volume
LV: left ventricular/ventricle
LA: left atrial/atrium
MAP: mean systemic arterial pressure
MCS: mechanical circulatory support
MPAP: mean pulmonary arterial pressure
PVA: Area of the PV loop/pressure-volume area
RA: right atrial/atrium
RV: right ventricular/ventricle
SW: stroke work
UPE: unavailable potential energy
V-A ECMO: veno-arterial extracorporeal membrane oxygenation

