## Supplementary Material for "Why does left ventricular distension usually not occur in veno-arterial extracorporeal membrane oxygenation?"

### Governing equations of the model

Left atrium and left ventricle:

$$\frac{dP_{la}}{dt} = \frac{1}{C_{la}}\left(\frac{P_{pv} - P_{la}}{R_{pv}} - F_{i,l}\right)$$

$$P_{sys,lv} = \begin{cases} 0 & if \quad V_{lv} \leq V_{u,lv} \\ E_{max,lv}\left(V_{lv} - V_{u,lv}\right) & if\ V_{u,lv} \leq V_{lv} \leq \dfrac{E_{max,lv}V_{u,lv} + P_{lv,d}}{E_{max,lv} - E_{lv,d}} \\ E_{lv,d}V_{lv} + P_{lv,d} & if \quad V_{lv} \geq \dfrac{E_{max,lv}V_{u,lv} + P_{lv,d}}{E_{max,lv} - E_{lv,d}} \end{cases}$$

$$P_{dia,lv} = P_{0,lv}\left(e^{k_{E,lv}(V_{lv} - V_{u,lv})} - 1\right)$$

$$P_{max,lv}(t) = \phi(t)P_{sys,lv} + [1 - \phi(t)]P_{dia,lv}$$

$$\phi(t) = \begin{cases} sin^2\left(\pi\dfrac{T}{T_{sys}}u(t)\right) & \text{if} \quad 0 \leq u \leq \dfrac{T_{sys}}{T} \\ 0 & \text{if} \quad \dfrac{T_{sys}}{T} \leq u(t) \leq 1 \end{cases}$$

$$u(t) = frac\left[\int_{t_0}^{t} \frac{1}{T(\tau)} d\tau + u(t_0)\right]$$

$$P_{lv} - P_{max,lv} + R_{lv}F_{o,l} = 0$$

$$\frac{dV_{lv}}{dt} = F_{i,l} - F_{o,l}\,, \quad where \quad F_{o,l} = \begin{cases} 0 & if \quad P_{lv} \leq P_{sa} \\ \dfrac{P_{lv} - P_{sa}}{R_{ao}} & if \quad P_{lv} > P_{sa} \end{cases}$$

$$F_{i,l} = \begin{cases} 0 & if \quad P_{la} \leq P_{lv} \\ \dfrac{P_{la} - P_{lv}}{R_{mv}} & if \quad P_{la} > P_{lv} \end{cases}$$

Systemic circulation:

$$\frac{dP_{sa}}{dt} = \frac{1}{C_{sa}}\left(F_{o,l} + F_{ec} - F_{sa} - \frac{P_{sa} - P_{art,c}}{R_{art,c}}\right)$$

$$\frac{dF_{sa}}{dt} = \frac{1}{L_{sa}}\left(P_{sa} - P_{sp} - R_{sa}.F_{sa}\right)$$

$$\frac{dP_{sp}}{dt} = \frac{dP_{ep}}{dt} = \frac{1}{C_{sp} + C_{ep}}\left(F_{sa} - \frac{P_{sp} - P_{sv}}{R_{sp}} - \frac{P_{sp} - P_{ev}}{R_{ep}}\right)$$

$$\frac{dP_{ev}}{dt} = \frac{1}{C_{ev}}\left(\frac{P_{sp} - P_{ev}}{R_{ep}} - \frac{P_{ev} - P_{ra}}{R_{ev}}\right)$$

$$P_{sv} = \frac{1}{C_{sv}}\big(V_t - C_{sa}P_{sa} - C_{art,c}P_{art,c} - V_{myo,c} - C_{ven,c}P_{ven,c} - (C_{sp} + C_{ep})P_{sp} - C_{ev}P_{ev} - C_{ra}P_{ra} - V_{rv} - C_{pa}P_{pa} - C_{pp}P_{pp} - C_{pv}P_{pv} - C_{la}P_{la} - V_{lv} - V_u\big)$$

$$V_u = V_{u,sa} + V_{u,sp} + V_{u,ep} + V_{u,sv} + V_{u,ev} + V_{u,ra} + V_{u,pa} + V_{u,pp} + V_{u,pv} + V_{u,la} + V_{u,lv} + V_{u,rv} + V_{myo,0}$$

Coronary circulation:

$$r_i = \left(\frac{3V_{lv}}{4\pi}\right)^{\frac{1}{3}}, \qquad r_0 = \left(\frac{3(V_{lv} + V_w)}{4\pi}\right)^{\frac{1}{3}}, \qquad \overline{r} = \left(\frac{3\left(V_{lv} + \frac{1}{3}V_w\right)}{4\pi}\right)^{\frac{1}{3}}$$

$$\overline{\lambda}_f = \left(\frac{V_{lv} + \frac{1}{3}V_w}{V_{u,lv} + \frac{1}{3}V_w}\right)^{\frac{1}{3}}, \qquad \overline{\lambda}_r = \frac{1}{\overline{\lambda}_f^{2}}, \qquad \overline{\sigma}_{mr} = \sigma_{mr,0}\left(e^{c_r(\overline{\lambda}_r - 1)} - 1\right)$$

$$P_{im} = \overline{\sigma}_{mr} + \frac{r_o - \overline{r}}{r_o - r_i}P_{lv}, \qquad P_{myo,c} = \overline{P}_{im} + \frac{V_{myo,c} - V_{myo,0}}{C_{myo,c}}$$

$$\frac{dP_{art,c}}{dt} = \frac{1}{C_{art,c}}\left(\frac{P_{sa} - P_{art,c}}{R_{art,c}} - \frac{P_{art,c} - P_{myo,c}}{R_{myo,u}}\right)$$

$$\frac{dV_{myo,c}}{dt} = \frac{P_{art,c} - P_{myo,c}}{R_{myo,u}} - \frac{P_{myo,c} - P_{ven,c}}{R_{myo,d}}$$

$$\frac{dP_{ven,c}}{dt} = \frac{1}{C_{ven,c}}\left(\frac{P_{myo,c} - P_{ven,c}}{R_{myo,d}} - \frac{P_{ven,c} - P_{ra}}{R_{ven,c}}\right)$$

Right atrium and right ventricle:

$$\frac{dP_{ra}}{dt} = \frac{1}{C_{ra}}\left(\frac{P_{sv} - P_{ra}}{R_{sv}} + \frac{P_{ev} - P_{ra}}{R_{ev}} + \frac{P_{ven,c} - P_{ra}}{R_{ven,c}} - F_{ec} - F_{i,r}\right)$$

$$P_{sys,rv} = \begin{cases} 0 & if \quad V_{rv} \le V_{u,rv} \\ E_{max,rv}(V_{rv} - V_{u,rv}) & if \ V_{u,rv} \le V_{rv} \le \dfrac{E_{max,rv}V_{u,rv} + P_{rv,d}}{E_{max,rv} - E_{rv,d}} \\ E_{rv,d}V_{rv} + P_{rv,d} & if \quad V_{lv} \ge \dfrac{E_{max,rv}V_{u,rv} + P_{rv,d}}{E_{max,rv} - E_{rv,d}} \end{cases}$$

$$P_{dia,rv} = P_{0,rv}\left(e^{k_{E,rv}(V_{rv}-V_{u,rv})} - 1\right)$$

$$P_{max,rv}(t) = \phi(t)P_{sys,rv} + [1 - \phi(t)]P_{dia,rv}$$

$$\phi(t) = \begin{cases} sin^2\left(\pi \frac{T}{T_{sys}} u(t)\right) & if \quad 0 \leq u \leq \frac{T_{sys}}{T} \\ 0 & if \quad \frac{T_{sys}}{T} \leq u(t) \leq 1 \end{cases}$$

$$u(t) = frac\left[\int_{t_0}^{t} \frac{1}{T(\tau)} d\tau + u(t_0)\right]$$

$$P_{rv} - P_{max,rv} + R_{rv}F_{o,r} = 0$$

$$\frac{dV_{rv}}{dt} = F_{i,r} - F_{o,r}\,, \quad where \quad F_{o,r} = \begin{cases} 0 & if \quad P_{rv} \leq P_{pa} \\ \frac{P_{rv} - P_{ra}}{R_{pu}} & if \quad P_{rv} > P_{pa} \end{cases}$$

$$F_{i,r} = \begin{cases} 0 & if \quad P_{ra} \leq P_{rv} \\ \frac{P_{ra} - P_{rv}}{R_{tv}} & if \quad P_{ra} > P_{rv} \end{cases}$$

Pulmonary circulation:

$$\frac{dP_{pa}}{dt} = \frac{1}{C_{pa}}\left(F_{o,r} - F_{p,a}\right)$$

$$\frac{dF_{pa}}{dt} = \frac{1}{L_{pa}}\left(P_{pa} - P_{pp} - R_{pa}.F_{pa}\right)$$

$$\frac{dP_{pp}}{dt} = \frac{1}{C_{pp}}\left(F_{pa} - \frac{P_{pp} - P_{pv}}{R_{pp}}\right)$$

$$\frac{dP_{pv}}{dt} = \frac{1}{C_{pv}}\left(\frac{P_{pp} - P_{pv}}{R_{pp}} - \frac{P_{pv} - P_{la}}{R_{pv}}\right)$$

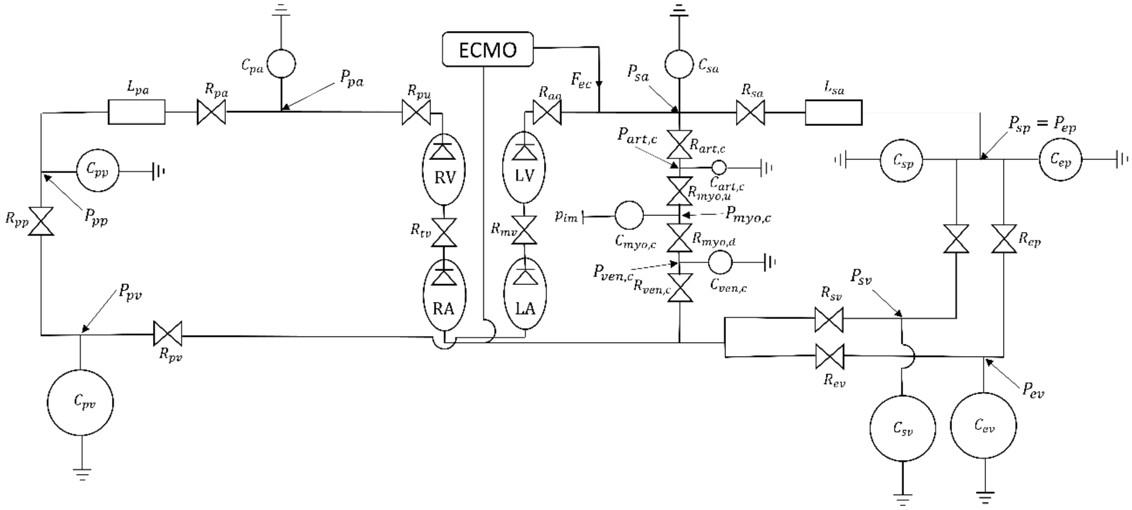


Figure S1: Schematic of the model

### Description of relevant pressure, volume, and flow variables used in the governing equations

Left atrial pressure - $P_{la}$

Left ventricular pressure - $P_{lv}$

Left ventricular volume - $V_{lv}$

Systemic arterial pressure - $P_{sa}$

Pulmonary arterial pressure - $P_{pa}$

Coronary blood flow rate - $\frac{P_{sa} - P_{art,c}}{R_{art,c}}$

Mean coronary blood flow rate ($Q_c$) - $\frac{1}{5T}\int_{t-5T}^{t} \frac{P_{sa} - P_{art,c}}{R_{art,c}} dt$

### Parameter values

Left atrium and left ventricle:

| | |
|---|---|
| $V_{u,la} = 25$ ml | $V_{u,lv} = 16.77$ ml |
| $C_{la} = 19.23$ ml/mmHg | $E^{n}_{max,lv} = 2.95$ mmHg/ml |
| $R_{mv} = 2.5 \times 10^{-3}$ mmHg/(ml/s) | $E_{lv,d} = -0.59$ mmHg/ml |
| $P_{0,\mathrm{lv}} = 1.5$ mmHg | $P_{lv,d} = 400$ mmHg |
| $k_{E,\mathrm{lv}} = 0.0182$ 1/ml | $R_{ao} = 6.0 \times 10^{-4}$ mmHg/ml.s |
| $k_{R,lv} = 3.75 \times 10^{-4}$ s/ml | $G_{P^{E}_{lv,d}} = 93.22$ ml |

Systemic circulation:

| Compliance (ml/mmHg) | Resistance (mmHg/(ml/s)) | Inertance (mmHg.ml/s$^2$) |
|---|---|---|
| $C_{sa} = 0.28$ | $R_{sa} = 0.06$ | $L_{sa} = 0.22 \times 10^{-3}$ |
| $C_{sp} = 1.44$ | $R_{sp} = 3.31$ | |
| $C_{ep} = 1.16$ | $R_{ep} = 1.41$ | |
| $C_{sv} = 61.11$ | $R_{sv} = 0.04$ | |
| $C_{ev} = 50.00$ | $R_{ev} = 0.02$ | |

Coronary circulation:

| Compliance (ml/mmHg) | Resistance (mmHg/(ml/s)) | Other parameters |
|---|---|---|
| $C_{art,c} = 0.004$ | $R_{art,c} = 5.25$ | $V_{myo,0} = 7$ ml |
| $C_{myo,c} = 0.187$ | $R_{myo,u} = 6.75$ | $V_w = 200\ ml$ |
| $C_{ven,c} = 0.093$ | $R_{myo,d} = 6.75$ | $c_r = 9$ |
| | $R_{ven,c} = 1.50$ | $\sigma_{mr,0} = 1.5 \times 10^{-3}$ mmHg |

Right atrium and right ventricle:

| | |
|---|---|
| $V_{u,ra} = 25$ ml | $V_{u,rv} = 40.8$ ml |
| $C_{ra} = 31.25$ ml/mmHg | $E^{n}_{max,rv} = 1.75$ mmHg/ml |
| $R_{tv} = 2.5 \times 10^{-3}$mmHg/ml. s | $E_{d,rv} = -0.35$ mmHg/ml |
| $P_{0,rv} = 1.5$ mmHg | $P_{d,rv} = 200$ mmHg |
| $k_{E,rv} = 0.0154$ 1/ml | $R_{pu} = 2.3 \times 10^{-4}$ mmHg/ml.s |
| $k_{R,rv} = 1.40 \times 10^{-4}$ s/ml | $G_{P^{E}_{rv,d}} = 71.43$ ml |

Pulmonary circulation:

| Compliance (ml/mmHg) | Resistance (mmHg/(ml/s)) | Inertance (mmHg.ml/s$^2$) |
|---|---|---|
| $C_{pa} = 0.76$ | $R_{pa} = 0.0230$ | $L_{pa} = 0.18 \times 10^{-3}$ |
| $C_{pp} = 5.80$ | $R_{pp} = 0.0894$ | |
| $C_{pv} = 25.37$ | $R_{pv} = 0.0056$ | |

Unstressed volumes (ml):

| | | | |
|---|---|---|---|
| $V_{u,sa} = 0$ | $V_{u,sp} = 274.4$ | $V_{u,ep} = 336.6$ | $V_{u,sv} = 1121$ |
| $V_{u,ev} = 1375$ | $V_{u,pa} = 0$ | $V_{u,pp} = 123$ | $V_{u,pv} = 120$ |

Other cardiovascular parameters

| | | |
|---|---|---|
| $T = 0.833s$ | $T_{sys} = 0.491s$ | $V_t = 5300$ ml |

Coronary-ventricular interaction parameters:

| Parameter | $Normal\ LV$ | $Moderate\ LV\ failure$ | $Severe\ LV\ failure$ |
|---|---|---|---|
| $E^0_{max,lv}$ (mmHg/ml) | 2.95 | 1.77 | 0.59 |
| $Q^0_{cor}$ (ml/min) | 277.0 | 242.5 | 140.8 |
| $Q_{c,max}$ (ml/min) | 300 | 300 | 300 |
| $Q_{c,min}$ (ml/min) | 100 | 100 | 100 |
| $G_{c,E_{lv}}$ ((mmHg/ml)/(ml/min)) | $3.0\ \times 10^{-3}$ | $3.0\ \times 10^{-3}$ | $3.0\ \times 10^{-3}$ |

### *Influence of $Q_{c,max}$ on the hemodynamic response*

$Q_{c,max}$ (the maximal coronary flow rate above which, $E_{max,lv}$ no longer changes with changes in $Q_c$) affects the hemodynamics response on V-A ECMO support, by limiting the maximum $E_{max,lv}$ an LV can achieve through improved coronary perfusion. Figure S2 presents the variation of hemodynamic variables with ECMO flow rate for different $Q_{c,max}$. A slightly larger reduction in LVEDV and LVESV occurred for higher $Q_{c,max}$. The improvement in LVEF was slightly higher for a higher $Q_{c,max}$. LVSW increased slightly as the ECMO flow rate increased, until it reached a maximum, and then decreased with a further increase in the ECMO flow rate. LVUPE increased almost linearly with ECMO flow rate nearly independent of the value of $Q_{c,max}$. LVPVA increased with increasing ECMO flow rate, again, a significant fraction of this increase was due to an increase in LVSW. Overall, the effects of changes in $Q_{c,max}$ were relatively modest.

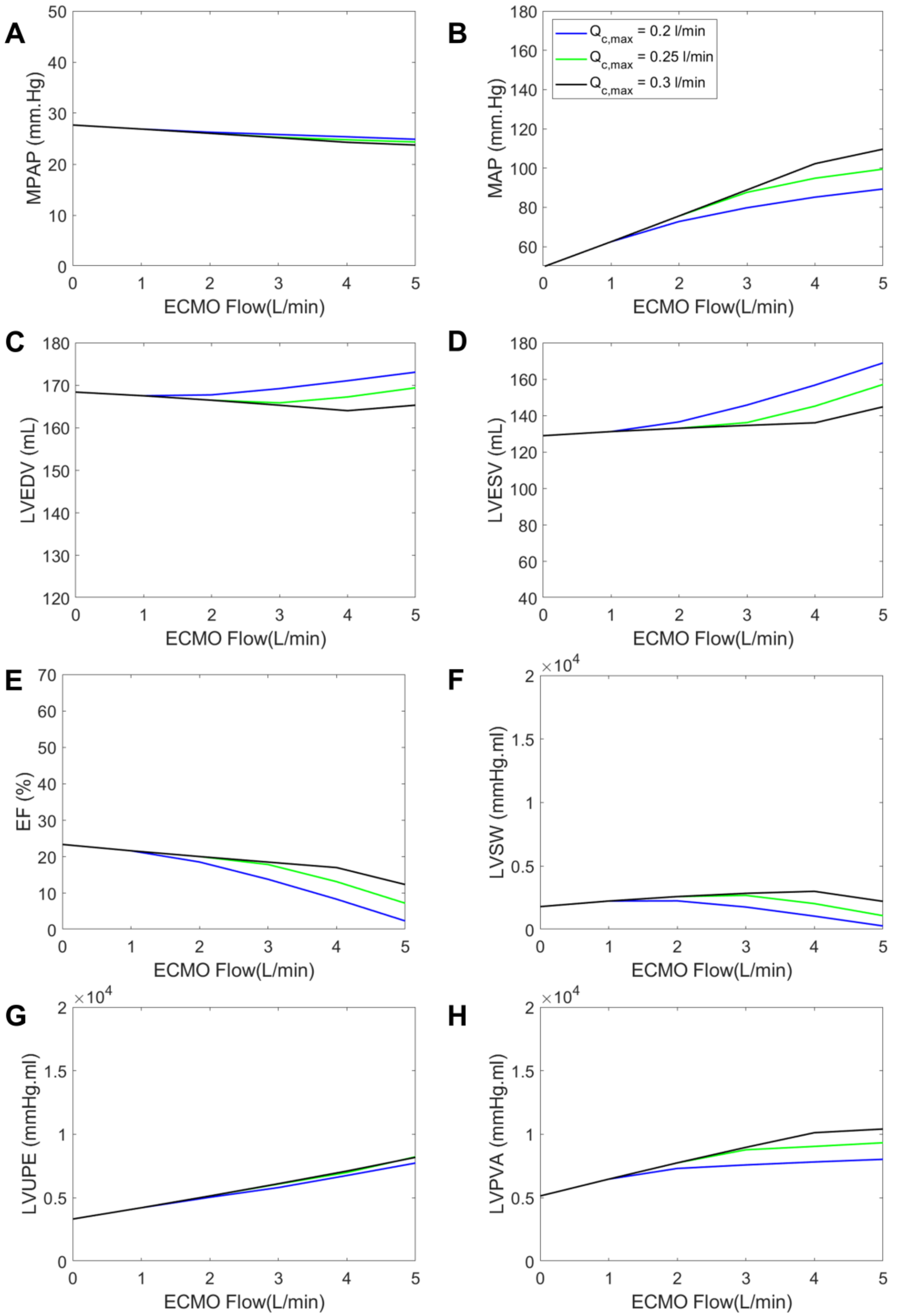
A
B
C
D
E
F
G
H
MPAP (mm.Hg)
MAP (mm.Hg)
LVEDV (mL)
LVESV (mL)
EF (%)
LVSW (mmHg.ml)
LVUPE (mmHg.ml)
LVPVA (mmHg.ml)
ECMO Flow(L/min)
Qc,max = 0.2 l/min
Qc,max = 0.25 l/min
Qc,max = 0.3 l/min
×10^4

**Figure S2:** Variation of hemodynamic variables with ECMO flow rate for different $Q_{c,max}$ in the case of severe left ventricular failure $E^{0}_{max,lv} = 0.2E^{n}_{max,lv}$. Legend represents the value of $Q_{c,max}$ in L/min. The value of $G_{c,E_{lv}}$ is fixed at $3 \times 10^{-3}$ $(mmHg/ml)/(ml/min)$. MAP – mean systemic arterial pressure; MPAP – mean pulmonary arterial pressure; LV – left ventricle; EDV - end-diastolic volume; ESV - end-systolic volume; EF – LV ejection fraction; SW – stroke work; UPE – unavailable potential energy;PVA – pressure-volume area.
